# Use of semaglutide and risk of non-arteritic anterior ischemic optic neuropathy: A Danish–Norwegian cohort study

**DOI:** 10.1101/2024.12.09.24318574

**Authors:** Emma Simonsen, Lars Christian Lund, Martin Thomsen Ernst, Vidar Hjellvik, Laszlo Hegedüs, Steffen Hamann, Øystein Kalsnes Jørstad, Hanne Løvdal Gulseth, Øystein Karlstad, Anton Pottegård

**Author notes:** **Correspondence,** Anton Pottegård, Professor, Clinical Pharmacology, Pharmacy and Environmental Medicine, University of Southern Denmark, Campusvej 55, DK-5230 Odense M, Denmark.

## Abstract

**Importance:** Recent findings have raised concerns about an association between semaglutide, a glucagon-like peptide-1 receptor agonist (GLP-1RA), and non-arteritic anterior ischemic optic neuropathy (NAION).

**Objective:** To further investigate the putative association between semaglutide use and NAION.

**Design:** Bi-national active comparator new-user cohort study with propensity score weighing to adjust for confounding, fixed-effect meta-analysis, and a supplementary self-controlled analysis (symmetry analysis).

**Setting:** Data were obtained from national health registries in Denmark and Norway, including prescription registries, patient registries, and civil registration systems.

**Participants:** New users of semaglutide and sodium-glucose co-transporter 2 inhibitors (SGLT-2is) during 2018-2022 (Norway) and 2018-2024 (Denmark).

**Exposures:** Initiation of semaglutide and SGLT-2is for the management of type 2 diabetes was assessed in the main analysis. In the self-controlled design, exposure to semaglutide, regardless of indication, was investigated.

**Main Outcomes and Measures:** Incidence rates of NAION among semaglutide and SGLT-2is users, along with hazard ratios (HRs) and incidence rate differences (IRDs) with robust 95% confidence intervals. Estimates from Denmark and Norway were pooled using a fixed-effects model (inverse variance weighting). The supplementary self-controlled study returned sequence symmetry ratios (SRs).

**Results:** We identified 44,517 eligible users of semaglutide for the management of type 2 diabetes in Denmark and 16,860 in Norway. In total, we observed 32 NAION events among semaglutide users (24 in Denmark and 8 in Norway). This yielded an unadjusted incidence rate of NAION of 2.19/10,000 person-years among Danish semaglutide initiators, compared to 1.18 among SGLT-2i initiators. The corresponding unadjusted incidence rates in Norway were 2.90 and 0.92. After adjustment, we obtained a pooled HR of 2.81 (95% CI 1.67 to 4.75) and IRD of +1.41 (95% CI +0.53 to +2.29). Estimates were compatible in the individual countries, although with higher and less precise estimates in Norway (HR 7.25; 95% CI 2.34 to 22.4) compared to Denmark (HR 2.17; 95% CI 1.20 to 3.92). Results were consistent across sensitivity and supplementary analysis, with a notably stronger association observed in a post hoc per-protocol analysis (HR 6.35; 95% CI 2.88 to 14.0). In the supplementary self-controlled study, SRs for NAION were 1.14 (95% CI 0.55 to 2.36) in Denmark and 2.67 (95% CI 0.91 to 8.99) in Norway.

**Conclusions and Relevance:** Our findings show an association between use of semaglutide for type 2 diabetes and risk of NAION. However, the absolute risk remains low.

**Key Points:** *Question:* Is there an association between semaglutide use and risk of non-arteritic anterior ischemic optic neuropathy (NAION)?

*Findings:* In this cohort study conducted in Denmark and Norway, use of semaglutide was associated with an increased risk of NAION; the pooled hazard ratio was 2.81 (95% confidence interval (CI) 1.67 to 4.75) and the incidence rate difference (absolute risk increase) was +1.41 (95% CI +0.53 to +2.29) NAION events per 10,000 person-years. The finding was consistent across sensitivity and supplementary analysis.

*Meaning:* Our study confirms that use of semaglutide is associated to an increased risk of NAION, but also that the excess absolute risk is low.

## Background

The glucagon-like peptide 1 receptor agonist (GLP-1RA) semaglutide has quickly become a key treatment for managing type 2 diabetes and obesity.^1^ Recent findings^2^ have raised concern about a potential association between semaglutide use and non-arteritic anterior ischemic optic neuropathy (NAION). NAION accounts for most cases of anterior ischemic optic neuropathy (AION), with the remaining cases classified as arteritic AION (AAION) due to giant cell arteritis.^3^ NAION is a major cause of vision loss in adults and is the second most frequent form of optic neuropathy after glaucoma.^4^ NAION presents as sudden, isolated, painless, monocular vision loss and optic disc edema. The vision loss is usually irreversible and there is no treatment. Given the serious nature of this potential adverse effect of semaglutide, we leveraged the nationwide Danish and Norwegian health registries to further investigate this association. Specifically, we performed a cohort study comparing the risk of NAION among individuals with type 2 diabetes using semaglutide compared to those using sodium-glucose co-transporter 2 inhibitors (SGLT-2is), and a self-controlled study examining the risk of NAION among semaglutide users, irrespective of indication of use.

## Methods

We assessed the occurrence of NAION among initiators of semaglutide and SGLT-2is in Denmark and Norway, using a common script approach. A target trial approach was applied, i.e., we designed the studies to mimic, as closely as possible, the corresponding hypothetical randomized controlled trial (RCT) that one would perform if the hypothesis that semaglutide increases NAION risk should be tested in a randomized design. SGLT-2is were selected as the most relevant comparator, as clinicians are recommended to either initiate SGLT-2is or GLP-1RAs for patients with type 2 diabetes and manifest cardiovascular disease or inadequate glycemic control with metformin.^5^ Hazard ratios (HRs) from Denmark and Norway were pooled using a meta-analysis approach. As a supplementary approach, we further evaluated the incidence of NAION after semaglutide initiation, regardless of indication, for each country in a self-controlled study using the sequence symmetry design.^6^

### Study transparency and reproducibility^7^

A study protocol was registered at the Real-World Evidence Registry before the commencement of any analyses. This protocol is available at https://osf.io/h3uv4, with amendments available at https://osf.io/9pdft. Expanded description of the methods applied, as well as all codes and definitions used in the study, can be found through these links. Data were used under license at both study sites and thus cannot be made publicly available. The source code used to perform data management and statistical analyses is made available at https://gitlab.sdu.dk/pharmacoepi/SemaglutideNAION. The manuscript is aligned with the Reporting of Studies Conducted using Observational Routinely Collected Health Data for pharmacoepidemiology (RECORD-PE) statement.^8^

### Data sources and linkage

Users of glucose-lowering therapies were identified using the Danish National Prescription Registry,^9^ covering all prescriptions filled at community pharmacies in Denmark since 1995, and the Norwegian Prescribed Drug Registry,^10^ covering all prescriptions filled at community pharmacies in Norway since 2004. Data on hospital admissions, including cases of the diagnosis “ischemic optic neuropathy” based on *International Classification of Diseases 10^th^ edition* (ICD-10) code were obtained from the Danish National Patient Registry,^11^ covering all hospital admissions in Denmark since 1977, and the Norwegian Patient Registry,^12^ covering all visits to specialist healthcare services including hospitals in Norway since 2008, as well as the Primary Care Registry,^12^ covering all visits to general practitioner physicians in Norway since 2017. Data on census were obtained from the Danish Civil Registration System^13^ and Statistics Norway,^14^ to account for death, censoring, and residence within Denmark and Norway, respectively. Data from laboratory analyses were exclusively available in Denmark and were obtained from the Danish Register of Laboratory Results.^15^ All data sources were linked using the unique personal civil registration number assigned to all Danish and Norwegian residents.^13^

### Study population and exposures

All new users of semaglutide and SGLT2-is in Denmark and Norway from the beginning of 2018 (year of market entry of semaglutide) through June of 2024 (Denmark) or May of 2022 (Norway) were identified, restricted to those using these drugs for the treatment of type 2 diabetes. In brief, for the Danish cohort, type 2 diabetes was defined as either having an HbA1c above 48 mmol/mol within two years before starting semaglutide or using metformin within 6 months before treatment initiation. In Norway, laboratory data were not available, and type 2 diabetes was therefore defined as having either (a) used metformin or sulfonylurea, (b) received a type 2 diabetes diagnosis from general practitioners in the primary healthcare sector, or (c) received a type 2 diabetes diagnosis in the specialist healthcare sector (somatic hospitals or outpatient private specialist practices contracted by the public regional health authorities) within one year before treatment initiation. We excluded everyone with a previous history of AION and those with migrations within two years before cohort entry. The primary exposure of interest was semaglutide initiation (first ever use), and the comparison cohort was initiators (also first ever use) of SGLT-2is. Initiators of semaglutide who had previously used SGLT-2is or other GLP-1RAs were excluded, as were initiators of SGLT-2is with a previous history of any GLP-1RA drug use.

#### Follow up and outcome

All patients were followed using an intention-to-treat approach, i.e., with no censoring upon treatment changes, switching or stopping, from the date of treatment initiation and for up to five years or until NAION diagnosis (main outcome), death, migration, or end of data availability (i.e., June of 2024 in Denmark and December of 2022 in Norway). NAION events were identified using the ICD-10 code H47.0c in Denmark and H47.0 in Norway, corresponding to AION, i.e., including both AAION and NAION. This was done under the assumption that the vast majority of outcome events would correspond to NAION, an assumption that was examined in a supplementary analysis. In Norway, both the main outcome and covariates were defined using data from the specialist health care sector, covering both contracted private-practicing specialists and hospitals. In Denmark, only diagnoses provided at hospitals were included.

#### Confounder adjustment

To reduce confounding, we used propensity scores to estimate the probability of receiving GLP-1RA treatment over SGLT-2i treatment based on a set of predefined characteristics. Propensity score models were created using logistic regression and included age, sex, calendar time, type of prescriber issuing the initial prescription (only available in Denmark), markers of diabetes severity, systemic risk factors for NAION, as well as specific comorbidities and co-medications. From the propensity scores, we calculated standardized mortality ratio (SMR) weights, assigning a weight of 1 to semaglutide initiators while SGLT-2i initiators were weighed according to their propensity score odds (propensity score divided by 1 minus their propensity score). This allowed us to create a pseudo-population of SGLT-2i users with characteristics similar to the semaglutide users, in turn allowing a balanced comparison of the two groups.^16^ The degree of comparability was evaluated using standardized mean differences (SMDs) with values less than 0.1 interpreted as a covariate being balanced.^17^

#### Missing information

To account for potential selection bias due to missing information in covariates, we used inverse probability (IP) weighting of complete cases.^18^ In brief, we estimated the probability of having complete information using logistic regression with independent variables being the same variables as included in the propensity score. Individuals with missing information received a weight of zero while complete cases were weighed as the proportion of complete cases over the probability of having complete information as per the model and using stabilized weights.

#### Analyses

We calculated the crude and adjusted (IP and SMR weighted) incidence rate of NAION in both the exposed and comparator cohort. The incidence rate differences were estimated using a Poisson model, using the log follow-up as offset. Additionally, we used Cox regression to estimate the crude (unweighted) and adjusted (weighted) hazard ratio (HR) associating semaglutide use with risk of NAION with robust 95% confidence intervals using a sandwich (Huber-White) estimator. HRs and IRDs from Denmark and Norway were pooled using inverse variance weighting (fixed-effects model).

#### Supplementary and sensitivity analyses

A series of supplementary and sensitivity analyses was carried out, some of which were pre-specified and some of which were post hoc analyses. More detailed reasoning for these analyses is provided in the protocol and its amendments (see above). First, we reduced the follow-up period to two years. Second, to mimic the Danish analyses, a sensitivity analysis was performed where the Norwegian analysis was restricted to outcomes registered in hospitals. Third, we applied a post hoc ‘per-protocol’ approach, censoring individuals from both the GLP-1RA and SGLT-2i cohorts if 180 days had passed with no filling of their index medication or upon filling of the opposite medication (i.e., GLP-1RA among SGLT-2i users and vice-versa). Fourth, due to skewness in the preference for SGLT-2i over GLP-1RA among patients with heart failure, we performed a post hoc analysis excluding individuals with a history of heart failure. Finally, also as a post hoc analysis, we evaluated the specificity of using the diagnosis code for AION to capture NAION events, by censoring all AION events where a diagnosis of giant cell arteritis was made for the same patient within two months before or after the AION event. This analysis could only be performed in Denmark.

### Self-controlled study

We conducted a sequence symmetry analysis (SSA), as described by Tsiropoulos et al.,^6^ to evaluate the incidence rate ratio of NAION^19^ following the initiation of semaglutide, regardless of indication. This included comparing NAION events during a one-year symmetric time window before and after starting semaglutide. If semaglutide was not associated with NAION, we expected NAION to occur at the same rate in both time periods. If semaglutide increased the risk of NAION, we expected more cases after starting semaglutide. The sequence ratio (SR) quantifies this comparison by dividing the number of post-treatment events by the number of pre-treatment events. SRs were adjusted for temporal trends in NAION occurrence,^6^ and 95% confidence intervals (CIs) were calculated using Jeffrey’s non-informative prior.^20^ In this analysis, all new users of semaglutide, irrespective of indication for use, were included up to one year prior to end of data availability (to allow for one year of follow-up). Further, for comparison, we performed the same analysis among new users of dapagliflozin and empagliflozin, the two most frequently prescribed SGLT-2is.

### Approvals

The Norwegian study was approved by the Regional Ethics Committee Sør-Øst B (28748, 2019/1175). Ethical approval is not required in Denmark for purely registry-based studies. The Danish study was registered at the repository of the University of Southern Denmark (11.570) and data access was granted by the Danish Health Data Authority (FSEID-6047).

## Results

In Denmark, we identified 58,336 initiators of semaglutide for the treatment of type 2 diabetes. Following restrictions, we included 44,517 eligible initiators of semaglutide as well as 84,814 initiators of SGLT-2is (**Figure S1**). Similarly, in Norway, we identified 21,023 initiators of semaglutide for the treatment of diabetes and ultimately included 16,860 semaglutide initiators and 34,153 initiators of SGLT-2is (**Figure S2**). For the combined cohort and before adjustment, semaglutide users were generally younger and had fewer comorbidities except for higher prevalence of obesity compared to users of SGLT-2is (**Table 1**). Characteristics of the two cohorts presented individually are available in **Table S1-2**.

**Table 1.** Baseline characteristics for new users of semaglutide and sodium-glucose co-transporter 2 inhibitors in Denmark and Norway combined before and after standardized mortality ratio weighting

|  | Unweighted cohort |  | SMR-weighted population |  |
| --- | --- | --- | --- | --- |
|  | Semaglutide<br>*n* = 60734 | SGLT-2is<br>*n* = 118967 | Semaglutide<br>*n* = 60887 | SGLT-2is<br>*n* = 60763 |
| Male sex | 33045 (54%) | 76338 (64%) | 33111 (54%) | 33215 (55%) |
| Age, years |  |  |  |  |
| <50 | 12906 (21%) | 13756 (12%) | 13037 (21%) | 13746 (23%) |
| 50-64 | 25100 (41%) | 41617 (35%) | 25174 (41%) | 24219 (40%) |
| 65-79 | 19617 (32%) | 50095 (42%) | 19582 (32%) | 19231 (32%) |
| 80+ | 3111 (5.1%) | 13499 (11%) | 3094 (5.1%) | 3568 (5.9%) |
| Diagnoses |  |  |  |  |
| Cerebrovascular disease | 3337 (5.5%) | 8535 (7.2%) | 3308 (5.4%) | 3397 (5.6%) |
| Heart failure | 2629 (4.3%) | 12564 (11%) | 2607 (4.3%) | 2670 (4.4%) |
| Obesity | 13086 (22%) | 11726 (9.9%) | 12985 (21%) | 13186 (22%) |
| Ischemic heart disease | 8146 (13%) | 24603 (21%) | 8080 (13%) | 8174 (13%) |
| Neurological complications | 2844 (4.7%) | 5404 (4.5%) | 2825 (4.6%) | 2946 (4.8%) |
| Peripheral vascular disease | 3300 (5.4%) | 8000 (6.7%) | 3277 (5.4%) | 3417 (5.6%) |
| Markers of alcohol abuse | 1827 (3%) | 3400 (2.9%) | 1813 (3%) | 1833 (3%) |
| Markers of smoking | 1727 (2.8%) | 2992 (2.5%) | 1710 (2.8%) | 1739 (2.9%) |
| Renal complications | 8917 (15%) | 21319 (18%) | 8835 (15%) | 9227 (15%) |
| Eye complications | 5224 (8.6%) | 10580 (8.9%) | 5192 (8.5%) | 5359 (8.8%) |
| Medications |  |  |  |  |
| Statins | 37875 (62%) | 82375 (69%) | 37551.18 (62%) | 37899 (62%) |
| Anticoagulants | 6148 (10%) | 17203 (14%) | 6099.95 (10%) | 6106 (10%) |
| Antiplatelets | 14753 (24%) | 38064 (32%) | 14625 (24%) | 14804 (24%) |
| ACEi/ARB | 36276 (60%) | 75861 (64%) | 35991 (59%) | 36155 (60%) |
| Amiodaron | 276 (0.45%) | 1085 (0.91%) | 273 (0.45%) | 275 (0.45%) |
| Phosphodiesterase-5 inhibitors * | 2989 (6.8%) | 6106 (7.2%) | 2951 (6.7%) | 2929 (6.6%) |
| Prescriber type * |  |  |  |  |
| GP | 38017 (87%) | 70860 (84%) | 38135 (63%) | 38409 (63%) |
| Hospital | 4648 (11%) | 12520 (15%) | 4658 (7.7%) | 4465 (7.3%) |
| Private practices | 343 (0.78%) | 489 (0.58%) | 351 (0.58%) | 255 (0.42%) |
| Unknown | 866 (2%) | 945 (1.1%) | 882 (1.4%) | 947 (1.6%) |
| Year of initiation |  |  |  |  |
| 2018 | 1091 (1.8%) | 6445 (5.4%) | 1073 (1.8%) | 1400 (2.3%) |
| 2019 | 6513 (11%) | 19030 (16%) | 6407 (11%) | 5829 (9.6%) |
| 2020 | 10324 (17%) | 21078 (18%) | 10236 (17%) | 11263 (19%) |
| 2021 | 16897 (28%) | 25088 (21%) | 16810 (28%) | 18433 (30%) |
| 2022 | 14098 (23%) | 20471 (17%) | 13994 (23%) | 13688 (23%) |
| 2023 * | 10616 (24%) | 17594 (21%) | 10541 (24%) | 7384 (17%) |
| 2024 * | 1838 (4.2%) | 9261 (11%) | 1826 (4.1%) | 2766 (6.3%) |

\* Data only available in Denmark

### Main analysis

A total of 32 NAION events were observed among semaglutide users (24 in Denmark and 8 in Norway). This yielded an unadjusted incidence rate of NAION of 2.19/10,000 person-years among Danish semaglutide initiators, compared to 1.18 among SGLT-2i initiators (**Table 2**). The corresponding incidence rates in Norway were 2.90 and 0.92 (**Table 2**).

**Table 2.** Rates of non-arteritic anterior ischemic optic neuropathy among semaglutide initiators compared to sodium-glucose co-transporter 2 inhibitors initiators and corresponding hazard ratios for Denmark and Norway and pooled incidence rate differences (measure of absolute risk) and hazard ratios (measure of relative risk) with up to five years of follow-up applying an ‘intention-to-treat’ approach.

|  | Incidence rate<br>per 10,000 py (events) |  | Hazard ratio<br>(95% CI) | Pooled IRD<br>per 10,000 py<br>(95% CI) | Pooled hazard<br>ratio (95% CI) |
| --- | --- | --- | --- | --- | --- |
|  | Semaglutide | SGLT-2is |  |  |  |
| Crude estimates |  |  |  |  |  |
| Denmark | 2.19 (24.0) | 1.18 (24.0) | 1.90 (1.08–3.35) | +1.18 (+0.29–+2.08) | 2.14 (1.31–3.52) |
| Norway | 2.90 (8.0) | 0.92 (7.0) | 3.21 (1.14–9.00) |  |  |
| Adjusted estimates <sup>a</sup> |  |  |  |  |  |
| Denmark | 2.18 (24.0) | 1.02 (11.6) | 2.17 (1.20–3.92) | +1.41 (+0.53–+2.29) | 2.81 (1.67–4.75) |
| Norway | 2.90 (8.0) | 0.40 (1.1) | 7.25 (2.34–22.4) |  |  |

Abbreviations: CI, confidence interval; IRD, incidence rate difference; SGLT-2is, sodium-glucose co-transporter 2 inhibitors; py, person-years
<sup>a</sup> adjusted estimates obtained in an standardized mortality ratio weighted pseudo-population

SMR weighting led to well-balanced patient characteristics, including, age, sex, and comorbidities, although with a slight residual imbalance in terms of year of treatment initiation (**Table 1** and **Table S1-2**). Estimates were compatible between the two countries, although with higher and less precise point estimates in Norway (adjusted HR 7.25; 95% CI 2.34 to 22.4) compared to Denmark (adjusted HR 2.17; 95% CI 1.20 to 3.92). The pooled adjusted HR was 2.81 (95% CI 1.67 to 4.75) with an incidence rate difference of +1.41/10,000 person-years (95% CI +0.53 to +2.29) (**Table 2**). For both countries, the cumulative incidence proportions for semaglutide and SGLT-2is started to diverge around 0.75 years after treatment initiation (**Figure 1**).

**Figure 1.**
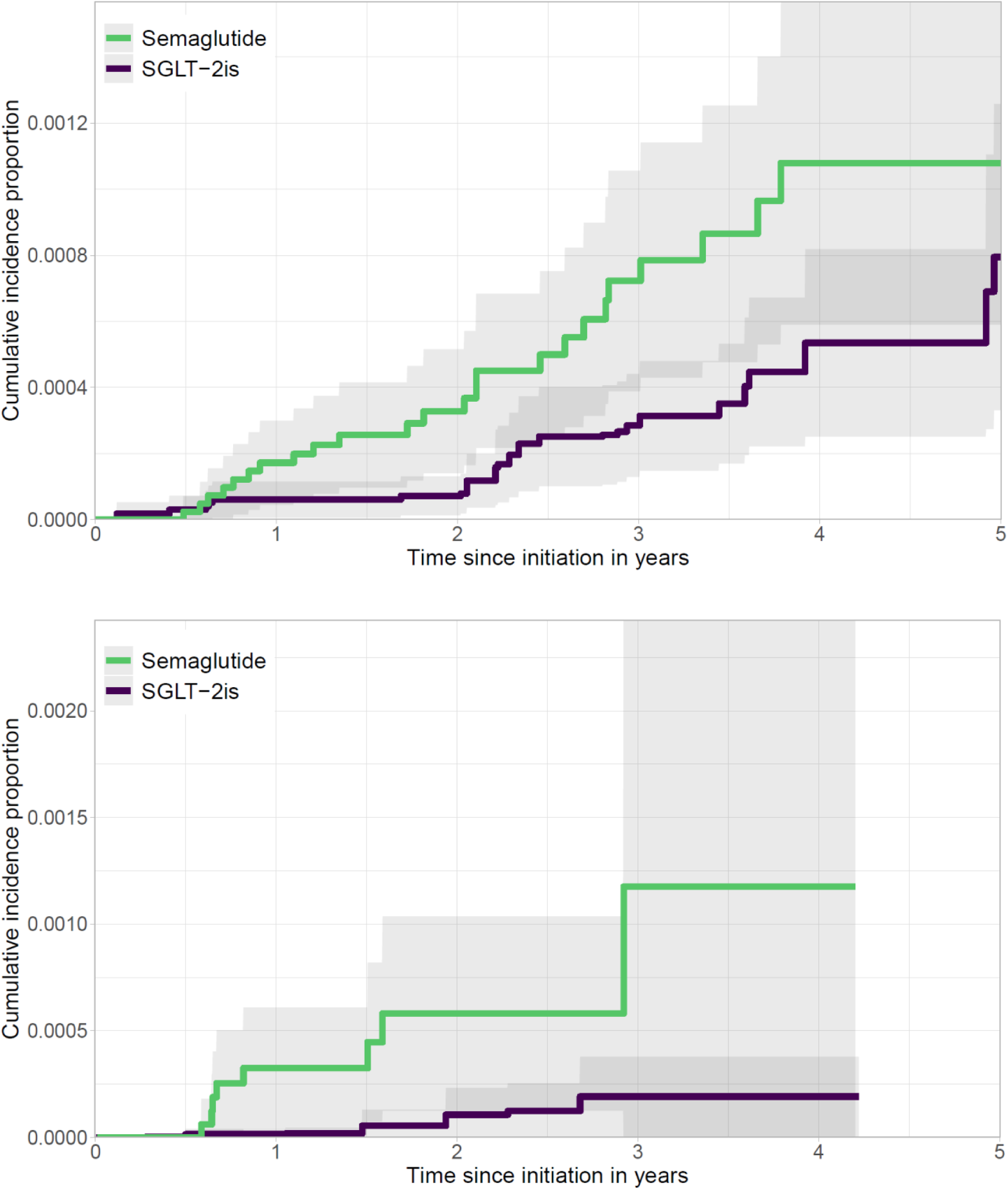
Weighted cumulative incidence proportion of non-arteritic anterior ischemic optic neuropathy outcomes with 95% confidence intervals (grey area) over time since initiation of semaglutide and sodium-glucose co-transporter 2 inhibitors initiators for Denmark (top panel) and Norway (bottom panel).

### Supplementary and sensitivity analyses

The results were generally consistent across supplementary and sensitivity analyses. A notably stronger risk estimate was observed in the post hoc ‘per-protocol’ analysis (HR 6.35; 95% CI 2.88 to 14.0) (**Table 3**). This increase was mainly explained by switching treatments, which was not taken into account in the ‘intention-to-treat’ approach of the main analysis. Specifically, of the 32 NAION events observed among SGLT-2i initiators in the main analysis, 17 occurred in users who had later switched to semaglutide after start of follow-up. The censoring of these individuals when they switched treatment in the ‘per-protocol’ analysis thus led to markedly lower rates of NAION in the comparison group, which in turn increased the observed HRs. Further, restricting the main analysis to a two-year follow-up period also resulted in higher but less precise risk estimates (HR 5.36; 95% CI 2.42 to 11.8) (**Table S3**). Limiting the Norwegian analysis to only hospital-registered NAION diagnoses yielded findings consistent with the main analysis (data not shown). Excluding individuals with heart failure from the analyses led to unchanged risk estimates (data not shown). Finally, the post hoc analysis of the Danish data, which censored outcome events with concomitant giant cell arteritis diagnosis, led to the removal of few outcomes (n<5), specifically among SGLT-2i users. This resulted in slightly stronger risk estimates (data not shown) and provided evidence that only NAION outcomes were observed among users of semaglutide in Denmark.

**Table 3.** Rates of non-arteritic anterior ischemic optic neuropathy among semaglutide initiators compared to sodium-glucose co-transporter 2 inhibitors initiators for Denmark and Norway and pooled incidence rate differences (measure of absolute risk) and hazard ratios (measure of relative risk) with up to five years of follow-up and applying a ‘per-protocol’ approach.

|  | Incidence rate<br>per 10,000 py (events) |  | Hazard ratio<br>(95% CI) | Pooled IRD<br>per 10,000 py<br>(95% CI) | Pooled hazard<br>ratio (95% CI) |
| --- | --- | --- | --- | --- | --- |
|  | Semaglutide | SGLT-2is |  |  |  |
| Crude estimates |  |  |  |  |  |
| Denmark | 2.88 (22.0) | 0.57 (7.0) | 4.95 (2.12–11.6) | +2.40 (+1.23–+3.59) | 5.23 (2.46–11.1) |
| Norway | 3.41 (<5*) | 0.47 (<5*) | 6.43 (1.24–33.3) |  |  |
| Adjusted estimates <sup>a</sup> |  |  |  |  |  |
| Denmark | 2.87 (22.0) | 0.54 (3.6) | 5.10 (2.14–12.2) | +2.46 (+1.30–+3.63) | 6.35 (2.88–14.0) |
| Norway | 3.41 (<5*) | 0.18 (<5*) | 18.1 (2.72–121) |  |  |

Abbreviations: CI, confidence interval; IRD, incidence rate difference; SGLT-2is, sodium-glucose co-transporter 2 inhibitors; py, person-years
<sup>a</sup> adjusted estimates obtained in an standardized mortality ratio weighted pseudo-population
\*Exact counts cannot be provided due to data privacy regulations

**Table 4.** Number of non-arteritic anterior ischemic optic neuropathy events before and after drug initiation and sequence ratios with 95% confidence intervals with the use of semaglutide (main exposure) or dapagliflozin and empagliflozin (sensitivity analyses).

|  | Sequences | SR<br>95% (CI) |
| --- | --- | --- |
| **Semaglutide** |  |  |
| Denmark | 15/14 | 1.14 (0.55–2.36) |
| Norway | 11/4 | 2.67 (0.91–8.99) |
| **Dapagliflozin and empagliflozin** |  |  |
| Denmark | 13/17 | 0.75 (0.36–1.52) |
| Norway | 11/14 | 0.77 (0.34–1.68) |

Abbreviations: CI, confidence interval; SR, symmetry ratio

### Self-controlled study

In the self-controlled study, we identified 221,636 individuals in Denmark and 38,019 individuals in Norway who initiated semaglutide, irrespective of indication. SR for NAION within one year was 1.14 (95% CI 0.55 to 2.36) in Denmark and 2.67 (95% CI 0.91 to 8.99) in Norway. Among initiators of dapagliflozin and empagliflozin (SGLT-2is), we found an SR of 0.75 (95% CI 0.36 to 1.52) in Denmark and 0.77 (95% CI 0.34 to 1.68) in Norway. During the one-year follow-up, less than five NAION events were seen among users of semaglutide for the treatment of obesity (data only available in Denmark).

### Post hoc descriptive analysis

To further explore the occurrence of NAION among users of semaglutide for obesity, we conducted an descriptive post hoc analysis of the Danish data. Here, we identified all users of semaglutide for the treatment of obesity (n = 156,937; median age 50 [IQR 40 to 58]) and followed them from treatment initiation until end of data availability (median follow-up 1.05 years; IQR 0.55 to 1.39). In this cohort, we observed eight events, corresponding to an incidence rate of 0.53 (95% CI 0.17 to 0.89).

## Discussion

In this population-based Danish–Norwegian cohort study, we observed an increased risk of NAION among users of semaglutide for type 2 diabetes compared to users of SGLT-2is. This finding was consistent across a range of sensitivity and supplementary analyses. However, the absolute risk of NAION remained low. Analyses of the risk of NAION with use of semaglutide for obesity were inconclusive.

The main strengths of the present analysis are the bi-national capture of all semaglutide users and their NAION events, the use of relevant active comparators, and the availability of high-quality healthcare registry data from two countries. The study also has several limitations. First, the relatively low number of NAION events hindered detailed subgroup analyses, limiting our understanding of how various characteristics (e.g., age, sex, and diabetes severity) may influence the risk of NAION. Second, the validity of AION diagnostic codes in our data is uncertain, as the diagnostic code used covers both the arteritic and non-arteritic forms of AION. However, AAION accounts for only 5-10% of AION cases and has a median age of 73 years and increases exponentially with age, with less than 10% occurring before the age of 60.^21,22^ Our cohorts had a median age of 65 years suggesting that the proportion of AAION cases in our data is likely at the lower end of the 5-10% range. Further, a post hoc analysis in the Danish cohort, which censored outcomes in patients with concurrent giant cell arteritis diagnosis (underlying any potential AAION events), resulted in slightly stronger risk estimates, further supporting the interpretation of the observed outcomes as true NAION events. Another concern could be the potential for underreporting AION cases referred to the hospitals. However, among ophthalmologists in Denmark and Norway, the prevailing clinical practice is that all suspected AION cases are referred to hospital, in particular, because of the importance of promptly distinguishing NAION from AAION requiring acute corticosteroid treatment. We therefore expect that very few, if any, AION cases are managed outside the hospital. This assumption is supported by data from Norway, where diagnoses from both contracted private practicing specialists and hospitals were available. In this dataset, we observed similar numbers of AION cases when restricting the analysis to hospital diagnoses, reinforcing the likelihood of comprehensive case capture in our cohort.

The safety signal associating use of semaglutide with risk of NAION was raised by Hathaway and colleagues,^2^ reporting a four-fold increased risk of developing NAION among patients with diabetes using semaglutide compared to patients with diabetes not using GLP-1RAs. Of note, this study was restricted to patients who had at some point (prior to inclusion or during follow-up) been referred for neuro-ophthalmological assessment. This raises concerns about selection bias, as also pointed out by others,^23^ since such referrals often are limited to individuals with more complex and severe conditions. The strong selection is illustrated by the fact that NAION, a relatively uncommon disease, was diagnosed in 8.9% of semaglutide users in the diabetes cohort. Whether this affected the relative risk estimates reported is unknown. Further, this selected population may not reflect the general demographic of patients with diabetes, potentially limiting generalizability.

To our knowledge, only two additional observational studies^24,25^ have assessed the risk of NAION among semaglutide users, but neither study found evidence of a statistically significant association. One of the cohort studies,^25^ which included data from 21 countries, observed a two-fold increase in NAION events among semaglutide users; however, the results of the study were not statistically significant, leading the authors to conclude that no such association was found. Of note, their findings are fully compatible with ours, which yielded similar estimates of risk, albeit with better statistical precision. Further, a meta-analysis of RCTs have investigated serious adverse effects of GLP-1RAs, including ischemic optic neuropathy.^26^ Sixty-nine trials were included in the meta-analysis, of which 64 did not report any case of NAION. In the remaining five trials, eight cases of ischemic optic neuropathy were observed in the GLP-1RA group and four cases in the comparator group. Of the eight cases reported in the GLP-1RA group, six occurred in patients treated with semaglutide, while liraglutide and dulaglutide accounted for one case each. Overall, GLP-1RA treatment was not associated with a significant increase in the risk of ischemic optic neuropathy. However, as NAION was not a prespecified safety outcome in these trials and the follow-up duration was limited, the meta-analysis was statistically underpowered to detect risk increases of the magnitudes reported in this paper.

Given the well-established effects of semaglutide in managing both diabetes^27^ and obesity,^28^ it is crucial to weigh the potential risk of NAION against the substantial therapeutic benefits of semaglutide. While the association observed for the use of semaglutide in type 2 diabetes represents a two-fold or higher relative risk increase, the absolute risk increase is limited, with as few as 1.4 to 2.5 (intention-to-treat and per-protocol estimates, respectively) excess NAION event per 10,000 years of follow-up among users of semaglutide for type 2 diabetes. Under the strong assumption of an elevated risk that is constant over time since treatment initiation, this corresponds to an absolute risk of 0.3-0.5% over 20 years use for type 2 diabetes. The analysis investigating the use of semaglutide for obesity and NAION risk yielded no evidence of such an association. However, since the study design required one year of available follow-up, only few individuals using semaglutide for obesity were included, and this analysis should thus be regarded as inconclusive rather than as providing evidence of the absence of an association. In the explorative post hoc analysis, we identified eight NAION events among Danish users of semaglutide for obesity. Although our findings thereby do not rule out the possibility of an increased risk of NAION when using semaglutide for obesity, the low number of observed events suggests that any potential risk is likely of limited absolute magnitude. Additional analyses with appropriate comparators or other study designs are needed to further investigate whether an increased risk of NAION is also evident among users of semaglutide for obesity and if so, the magnitude of this risk.

In conclusion, our findings support an association between use of semaglutide for type 2 diabetes and risk of NAION, with a more than two-fold increased hazard ratio. However, the absolute risk of NAION remains low among semaglutide users. Analyses of an association between semaglutide for obesity and NAION were inconclusive.

## Data Availability

All data produced in the present study are available upon reasonable request to the authors.

## Acknowledgements

The authors would like to acknowledge Peter Bjødstrup Jensen for validation of the analytical code used.

