## Supplementary Material for "Use of semaglutide and risk of non-arteritic anterior ischemic optic neuropathy: A Danish–Norwegian cohort study"


Emma Simonsen[1] MD
Lars Christian Lund[1] MD PhD
Martin Thomsen Ernst[1] MSc
Vidar Hjellvik[2] PhD
Laszlo Hegedüs[3,4] MD DMSc
Steffen Hamann[5,6] MD PhD
Øystein Kalsnes Jørstad[7] MD PhD
Hanne Løvdal Gulseth[2] MD PhD
Øystein Karlstad[2] MSc PhD
Anton Pottegård[1] MSc PhD DMSc

(1) Clinical Pharmacology, Pharmacy, and Environmental Medicine, Department of Public health, University of Southern Denmark, Odense, Denmark
(2) Department of Chronic Diseases, Norwegian Institute of Public Health, Oslo, Norway
(3) Department of Endocrinology, Odense University Hospital, Odense, Denmark
(4) Department of Regional Health Research, University of Southern Denmark, Odense, Denmark
(5) Department of Ophthalmology, Copenhagen University Hospital - Rigshospitalet, Copenhagen, Denmark
(6) Department of Clinical Medicine, University of Copenhagen, Copenhagen, Denmark
(7) Department of Ophthalmology, Oslo University Hospital, Oslo, Norway and Faculty of Medicine, University of Oslo, Oslo, Norway

**Correspondence**
Anton Pottegård
Professor
Clinical Pharmacology, Pharmacy
and Environmental Medicine
University of Southern Denmark
Campusvej 55
DK-5230 Odense M, Denmark

ORCID: 0000-0001-9314-5679

**Table S1** Baseline characteristics for new users of semaglutide and sodium-glucose co-transporter 2 inhibitors in Denmark before and after standardized mortality ratio weighting

| | **Unweighted cohort** | | | **SMR-weighted population** | | |
|---|---|---|---|---|---|---|
| | **Semaglutide** *n* = 44517 | **SGLT-2is** *n* = 84814 | **SMD** | **Semaglutide** *n* = 44027 | **SGLT-2is** *n* = 44076 | **SMD** |
| Male sex | 24283 (55) | 53943 (64) | 0.18 | 24012 (55) | 24079 (55) | <0.01 |
| Age, years | | | 0.39 | | | 0.05 |
| <50 | 9139 (21) | 9271 (11) | | 9045 (21) | 9648 (22) | |
| 50-64 | 18135 (41) | 28909 (34) | | 17930 (41) | 17411 (40) | |
| 65-79 | 14780 (33) | 36152 (43) | | 14619 (33) | 14232 (32) | |
| 80+ | 2463 (5.5) | 10482 (12) | | 2434 (5.5) | 2785 (6.3) | |
| Diagnoses | | | | | | |
| Cerebrovascular disease | 2499 (5.6) | 6170 (7.3) | 0.07 | 2470 (5.6) | 2518 (5.7) | <0.01 |
| Heart failure | 1781 (4.0) | 8440 (10) | 0.24 | 1759 (4.0) | 1787 (4.1) | <0.01 |
| Obesity | 9375 (21) | 8743 (10) | 0.30 | 9274 (21) | 9513 (22) | 0.01 |
| Ischemic heart disease | 5403 (12) | 15169 (18) | 0.16 | 5337 (12) | 5394 (12) | <0.01 |
| Neurological complications | 1928 (4.3) | 3603 (4.3) | <0.01 | 1909 (4.3) | 1957 (4.4) | 0.01 |
| Peripheral vascular disease | 2242 (5.0) | 5218 (6.2) | 0.05 | 2219 (5.0) | 2277 (5.2) | 0.01 |
| Markers of alcohol abuse | 1448 (3.3) | 2754 (3.3) | <0.01 | 1434 (3.3) | 1454 (3.3) | <0.01 |
| Markers of smoking | 1524 (3.4) | 2644 (3.1) | 0.02 | 1507 (3.4) | 1533 (3.5) | <0.01 |
| Renal complications | 7274 (16) | 18169 (21) | 0.13 | 71912 (16) | 7390 (17) | 0.01 |
| Eye complications | 2526 (5.7) | 4648 (5.5) | 0.01 | 2494 (5.7) | 2574 (5.8) | 0.01 |
| Medications | | | | | | |
| Statins | 28913 (65) | 61482 (72) | 0.16 | 28589 (65) | 28791 (65) | 0.01 |
| Anticoagulants | 4436 (10) | 12337 (15) | 0.14 | 4388 (10) | 4389 (10) | <0.01 |
| Antiplatelets | 10332 (23) | 25778 (30) | 0.16 | 10204 (23) | 10285 (23) | <0.01 |
| ACEi/ARB | 27201 (61) | 56125 (66) | 0.11 | 26916 (61) | 27020 (61) | <0.01 |
| Amiodaron | 207 (0.46) | 744 (0.88) | 0.05 | 204 (0.46) | 205 (0.47) | <0.01 |
| Phosphodiesterase-5 inhibitors | 2989 (6.7) | 6106 (7.2) | 0.02 | 2951 (6.7) | 2929 (6.7) | <0.01 |
| Prescriber type | | | 0.14 | | | 0.03 |
| GP | 38548 (87) | 70860 (84) | | 38135 (87) | 38409 (87) | |
| Hospital | 4715 (11) | 12520 (15) | | 4658. (11) | 4465 (10) | |
| Private practices | 355 (0.80) | 489 (0.58) | | 351 (0.80) | 255 (0.58) | |
| Unknown | 899 (2.0) | 945 (1.1) | | 882 (2.0) | 947 (2.2) | |
| Year of initiation | | | 0.31 | | | 0.21 |
| 2018 | 1085 (2.4) | 3875 (4.6) | | 1067 (2.4) | 1193 (2.7) | |
| 2019 | 4759 (11) | 10644 (13) | | 4653 (11) | 4397 (10) | |
| 2020 | 6730 (15) | 12016 (14) | | 6642 (15) | 7290 (17) | |
| 2021 | 8978 (20) | 14879 (18) | | 8891 (20) | 10738 (24) | |
| 2022 | 10511 (24) | 16545 (20) | | 10407 (24) | 10308 (23) | |
| 2023 | 10616 (24) | 17594 (21) | | 10541 (24) | 7384 (17) | |
| 2024 | 1838 (4.1) | 9261 (11) | | 1826 (4.2) | 2766 (6.3) | |

**Table S2** Baseline characteristics for new users of semaglutide and sodium-glucose co-transporter 2 inhibitors in Norway before and after standardized mortality ratio weighting

| | **Unweighted cohort** | | | **SMR-weighted population** | | |
|---|---|---|---|---|---|---|
| | **Semaglutide** *n* = 16860 | **SGLT-2is** *n* = 34153 | **SMD** | **Semaglutide** *n* = 16860.00 | **SGLT-2is** *n* = 16685.28 | **SMD** |
| Male sex | 9099 (54) | 22395 (66) | 0.24 | 9099 (54) | 9137 (55) | 0.02 |
| Age, years | | | 0.39 | | | 0.05 |
| <50 | 3992 (24) | 4485 (13) | | 3992 (24) | 4099 (25) | |
| 50-64 | 7244 (43) | 12708 (37) | | 7244 (43) | 6802 (41) | |
| 65-79 | 4964 (29) | 13943 (41) | | 4964 (29) | 5000 (30) | |
| 80+ | 660 (3.9) | 3017 (8.8) | | 660 (3.9) | 784 (4.7) | |
| Diagnoses | | | | | | |
| Cerebrovascular disease | 838 (5.0) | 2365 (6.9) | 0.08 | 838 (5.0) | 879 (5.3) | 0.01 |
| Heart failure | 848 (5.0) | 4124 (12) | 0.25 | 848 (5.0) | 883 (5.3) | 0.01 |
| Obesity | 3711 (22) | 2983 (8.7) | 0.37 | 3711 (22) | 3670 (22) | <0.01 |
| Ischemic heart disease | 2743 (16) | 9434 (28) | 0.28 | 2743 (17) | 2781 (17) | 0.01 |
| Neurological complications | 916 (5.4) | 1801 (5.3) | 0.01 | 916 (5.4) | 989 (5.9) | 0.02 |
| Peripheral vascular disease | 1058 (6.3) | 2782 (8.2) | 0.07 | 1058 (6.3) | 1140 (6.8) | 0.02 |
| Markers of alcohol abuse | 379 (2.3) | 646 (1.9) | 0.03 | 379 (2.3) | 380 (2.3) | <0.01 |
| Markers of smoking | 203 (1.2) | 348 (1.0) | 0.02 | 203 (1.2) | 207 (1.2) | <0.01 |
| Renal complications | 1643 (9.7) | 3150 (9.2) | 0.02 | 1643 (9.7) | 1836 (11) | 0.04 |
| Eye complications | 2698 (16) | 5932 (17) | 0.04 | 2698 (16) | 2785 (17) | 0.02 |
| Medications | | | | | | |
| Statins | 8962 (53) | 20893 (61) | 0.16 | 8962 (53) | 9108 (55) | 0.03 |
| Anticoagulants | 1712 (10) | 4866 (14) | 0.13 | 1712 (10) | 1717 (10) | <0.01 |
| Antiplatelets | 4421 (26) | 12286 (36) | 0.21 | 4421 (26) | 4520 (27) | 0.02 |
| ACEi/ARB | 9075 (54) | 19736 (58) | 0.08 | 9075 (54) | 9134 (55) | 0.02 |
| Amiodaron | 69 (0.41) | 341 (1.0) | 0.07 | 69 (0.41) | 70 (0.42) | <0.01 |
| Phosphodiesterase-5 inhibitors* | - | - | | - | - | |
| Year of initiation | | | 0.67 | | | 0.17 |
| 2018 | 6 (0.04) | 2570 (7.5) | | 6 (0.04) | 207 (1.2) | |
| 2019 | 1754 (10) | 8386 (25) | | 1754 (10) | 1432 (8.6) | |
| 2020 | 3594 (21) | 9062 (27) | | 3594 (21) | 3973 (24) | |
| 2021 | 7919 (47) | 10209 (30) | | 7919 (47) | 7696 (46) | |
| 2022 | 3587 (21) | 3926 (12) | | 3587 (21) | 3378 (20) | |

* Not available in Norwegian dataset

**Table S3** Rates of non-arteritic anterior ischemic optic neuropathy among semaglutide initiators compared to sodium-glucose co-transporter 2 inhibitors initiators for Denmark and Norway and pooled incidence rate differences (measure of absolute effect size) and hazard ratios (measure of relative effect size) with two-year follow-up (intention-to-treat approach)

| | **Incidence rate per 10,000 py (events)** | | **Hazard ratio (95% CI)** | **Pooled IRD per 10,000 py (95% CI)** | **Pooled hazard ratio (95% CI)** |
|---|---|---|---|---|---|
| | Semaglutide | SGLT-2is | | | |
| Crude estimates | | | | | |
| Denmark | 1.64 (12) | 0.47 (6) | 3.48 (1.31–9.28) | 1.32 (0.40–2.24) | 3.43 (1.63–7.25) |
| Norway | 2.91 (7) | 0.89 (5) | 3.37 (1.06–10.7) | | |
| Adjusted estimates [a] | | | | | |
| Denmark | 1.64 (12.0) | 0.39 (2.8) | 4.19 (1.53–11.5) | 1.47 (0.57–2.36) | 5.36 (2.42–11.8) |
| Norway | 2.91 (7.0) | 0.37 (0.9) | 7.98 (2.21–28.8) | | |

Abbreviations: CI, confidence interval; IRD, incidence rate difference; SGLT-2is, sodium-glucose co-transporter 2 inhibitors; py, person-years
[a] adjusted estimates obtained in an standardized mortality ratio weighted pseudo-population

**Supplementary Figure S1** Flowchart of cohort selection for Denmark

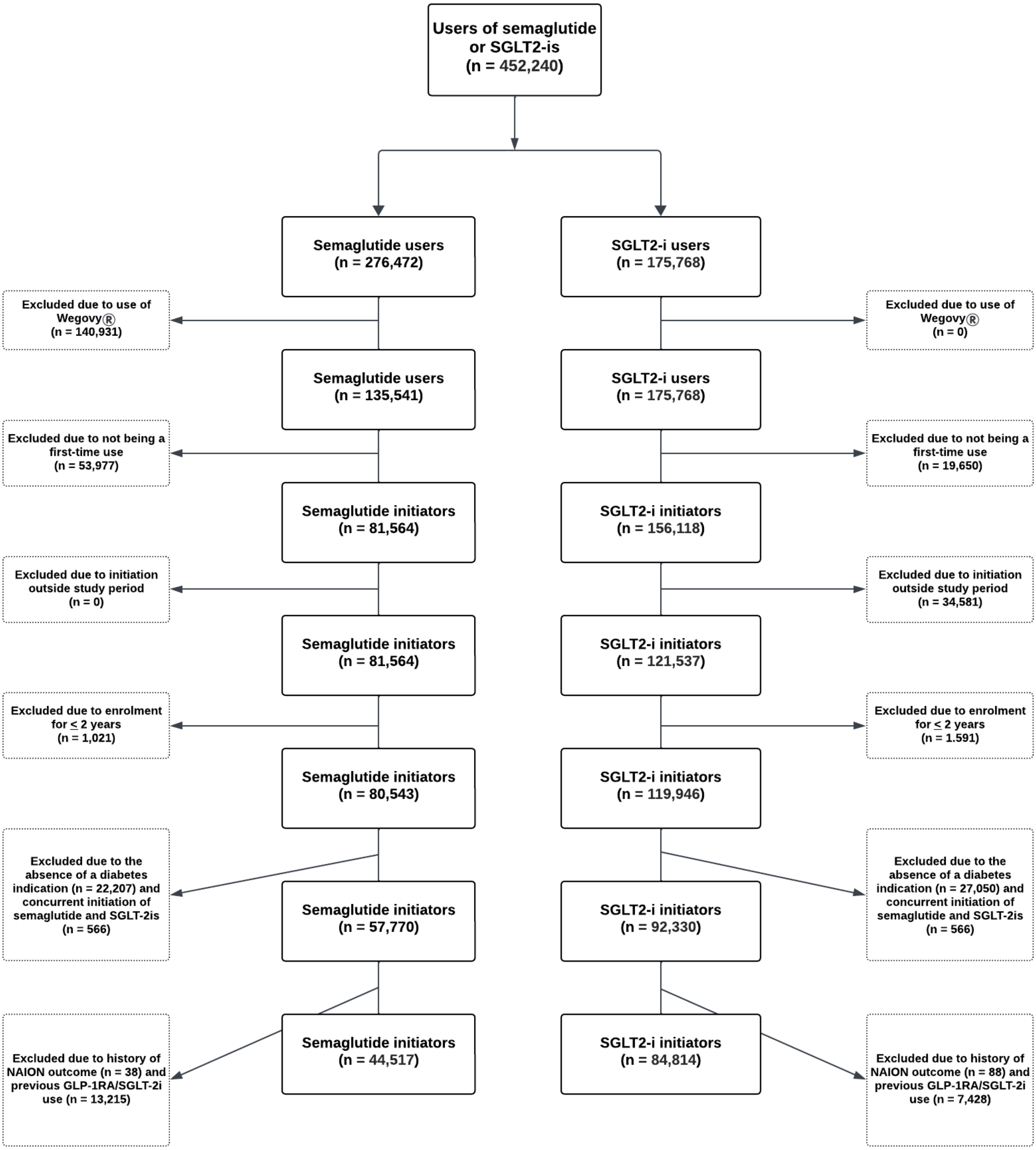

**Supplementary Figure S1** Flowchart of cohort selection for Norway

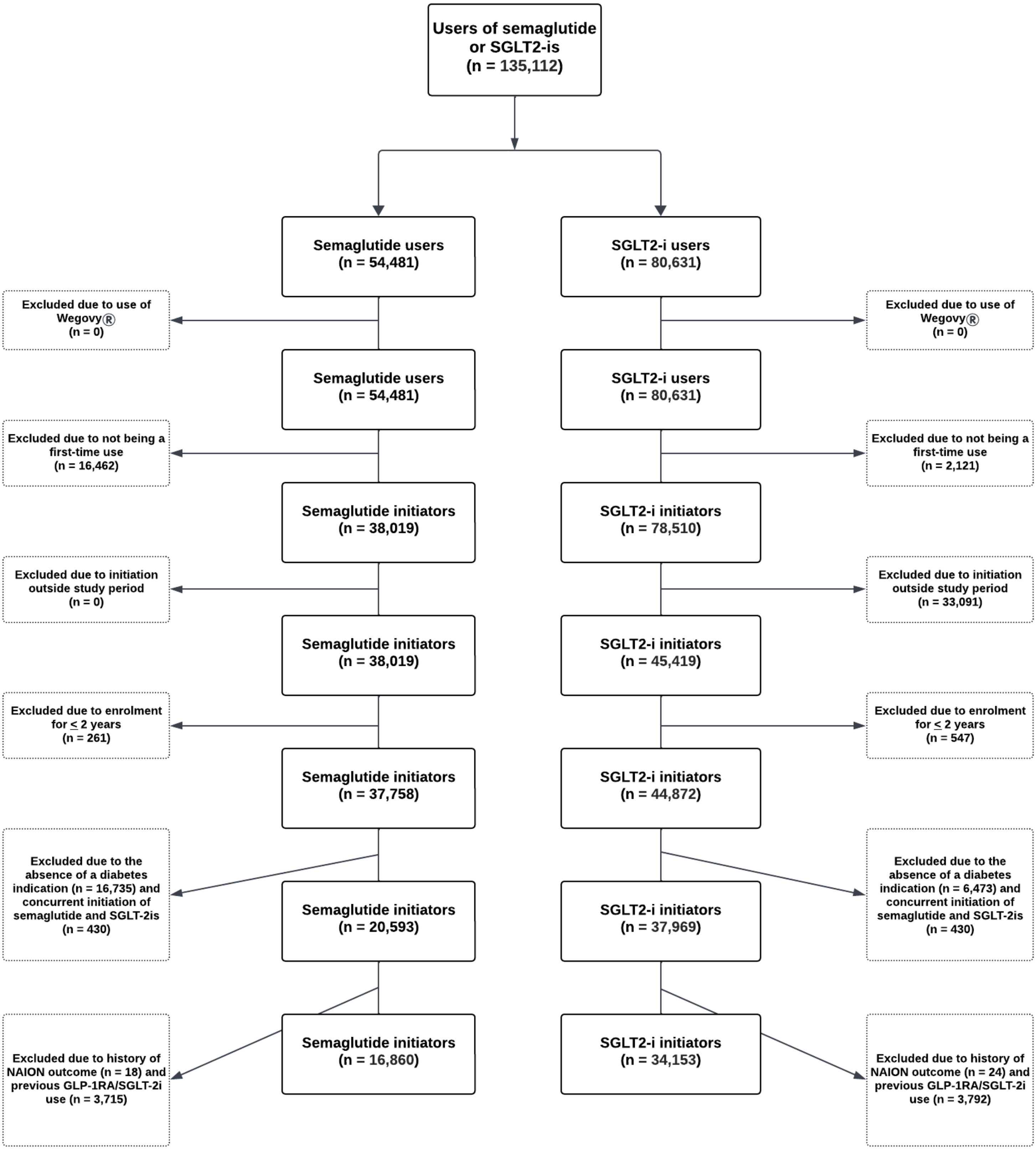
